# Using Over-the-Counter Retail Medication Sales to Detect and Track Influenza-Like Illnesses Including Novel Diseases

**DOI:** 10.1101/2025.11.07.25339792

**Authors:** Ye Ye, Jessi Espino, John M. Aronis, Harry Hochheiser, Marian G. Michaels, Gregory F. Cooper

## Abstract

Timely detection of emerging disease outbreaks is critical for effective public health response. Traditional surveillance systems often rely on clinical or laboratory-confirmed data, which may delay early detection. In this study, we evaluate the potential of the National Retail Data Monitor (NRDM), which tracks over-the-counter (OTC) health-related product sales, as a tool for public health surveillance. Using data from Allegheny County, Pennsylvania (2016–2021), we developed a probabilistic model that estimates daily influenza-like illness (ILI) activity based on purchasing patterns. To train the model, we applied an expectation-maximization (EM) algorithm to estimate the conditional probabilities of OTC product purchases given ILI or not ILI, which forms the basis of our model. To test the model, we used that model and daily OTC sales to estimate the number of purchases each day due to ILI. The estimated ILI counts showed moderately strong correlation (r = 0.66) with ILI counts derived from emergency departments (EDs) in the same region. We also monitor the extent to which our model predicts the data well; if the current prediction is poor, relative to historical predictions, it raises the prospect that there is an outbreak of an unusual disease in the population, perhaps even a novel disease. Our findings suggest that surveillance of OTC products can effectively identify unusual health-related activity and provide early insights of potential known and novel outbreaks through changes in consumer purchasing behavior.

## 1. Introduction

Detection of disease outbreaks is critical to public health surveillance, particularly in the early identification of emerging outbreaks. Traditional surveillance approaches, such as sentinel systems operated by the Centers for Disease Control and Prevention (CDC), rely on confirmed case reports from selected healthcare systems. While these methods provide accurate clinical insights, they can suffer from reporting delays, limiting their effectiveness for timely response. To address this gap, public health surveillance has evolved to incorporate diverse data streams, including electronic health records (EHRs) (Elkin, 2012; Ye, 2017), search engine query (Ginsberg, 2009), and social media (Jordan, 2018; Zhang, 2022) to support timely detection. EHR-based surveillance enables real-time insights from structured healthcare data but depends on integration with clinical systems. Search engine-based surveillance leverages aggregated query trends, as for example spikes in Google searches for influenza symptom, to estimate disease activity in near real time. Social-media-based surveillance mines user-generated content, such as symptom-related posts, to detect unusual patterns in health behavior.

An additional, underutilized approach involves analyzing over-the-counter (OTC) product sales, which can reflect self-treatment behaviors (Johnson 2005; Hogan 2006). Since these purchases often precede formal medical care, anomalies in OTC sales can signal emerging population-level health events. To support early outbreak detection nationwide, the National Retail Data Monitor (NRDM) (Wagner, 2003; Wagner, 2004) was launched in December 2002 as a novel public health surveillance system. Operated by the University of Pittsburgh, the NRDM has collected anonymous sales data on selected OTC healthcare products, such as antipyretics, cough and cold medications, and thermometers from more than 28,000 participating retail pharmacies, grocery stores, and mass merchandise outlets across the U.S.

NRDM was developed to enhance public health capacity for syndromic surveillance, enabling rapid detection of unusual health events through deviations in consumer purchasing behavior. More than 800 public health officials in 49 states, the District of Columbia, and U.S. territories have had access to NRDM through secure online portals (Espino, 2004; Tsui 2005), allowing them to track anomalies in consumer health behavior in near real-time. Since the launch of the NRDM, many studies have been conducted to enhance its functionality, including the development of fast spatial and space-time scan algorithms (Neill, 2005) and statistics (Sabhnani, 2005), the integration of user feedback mechanisms (Sabhnani, 2006), evaluation of policies for updating product categories (Hogan, 2005), and analysis of the correlation between OTC sales and estimated daily influenza-like illness in emergency departments (Villamarín, 2013), as well as the relationship between OTC pharmaceutical sales and patient volume at urgent care centers (Liu, 2013). Numerous studies have also validated the value of NRDM in complementing traditional clinical surveillance systems for investigations of gastrointestinal illness outbreaks — including those in Washoe County, Nevada (Chen, 2005), the San Francisco Bay Area (Kirian, 2010), and in New York City in the context of a massive power outage in August 2003 (Marx, 2006). NRDM data have also been used to help detect spikes in unintentional poisonings (Krenzelok, 2008), support syndromic surveillance during the San Diego wildfires in 2003 (Johnson, 2005), monitor sales of electrolyte products as early indicators of respiratory and diarrheal outbreaks in children (Hogan, 2003), and the data was a key component of New York City’s syndromic surveillance systems (Heffernan, 2004).

We previously developed ILI Tracker (Aronis 2024), a Bayesian surveillance method that uses emergency department (ED) data to estimate the daily frequency of multiple influenza-like illnesses, including potential novel diseases. As an illustrative example, retrospective analysis indicated that applying ILI Tracker to 2014 data would have enabled early detection of an enterovirus D68 outbreak in Allegheny County. Building on this foundation, we sought to determine whether pre-diagnostic, population-level signals—specifically, OTC product purchases—could enable even earlier outbreak detection. To this end, we developed and evaluated a similar probabilistic modeling system that uses NRDM data to detect outbreaks of known ILI diseases. The system also seeks to detect the onset of novel diseases by identifying statistically significant deviations in OTC product purchases relative to expected purchases for known disease models. We called this system *NRDM Tracker*. We evaluated NRDM Tracker on five years of real OTC data in Allegheny County. The results indicate that NRDM Tracker was able to detect both known and novel diseases, showing the system’s usefulness in public health surveillance.

## 2. Methods

### 2.1 Research data

We retrieved OTC product sales for Allegheny County, Pennsylvania (270 stores) from June 1, 2016 to December 30, 2021 from the NRDM database. We then divided the available NRDM OTC product categories into the seven categories shown in Table 1.

**Table 1.** Seven OTC product categories in NRDM.

| Category | Description |
| --- | --- |
| Cough or Cold | All cough and cold related medications, including pediatric and adult formulations of liquids and tablets, bronchial remedies, chest rubs, nasal sprays and throat lozenges. |
| Antifever Adult | All products intended for adults that are taken orally for the relief of fever |
| Antifever Pediatric | All products intended for children that are taken orally for the relief of fever |
| Baby or Child Electrolyte | Solutions of primarily electrolytes and glucose used for fluid replacement in children suffering from dehydration; includes products in liquid and freezer pop form |
| Thermometer | All thermometers, designed for both pediatric and adult use, including electronic and “ear” aka tympanic membrane thermometers used for health reasons; also includes non-intrusive fever detectors and replacement sheaths for electronic thermometers |
| Antidiarrheal | Products taken for the relief of diarrhea; all sizes and forms (tablets, liquids, etc.); includes bismuth (Pepto Bismol and like products), attapulgit, subsalicylate, and loperamide HCL products |
| Hydrocortisone product | Anti-inflammatory topical corticosteroids used to relieve itching rashes |

To support the method, we used for estimating parameters in the NRDM-Tracker model, we needed to estimate the prior probabilities of influenza-like illness (ILI) in the population (Section 2.3). To do so, we obtained daily counts of ED encounters associated with ILI-related viruses in the region, including adenovirus, rhinovirus/enterovirus, influenza, human metapneumovirus (HMPV), parainfluenza (PIV), and respiratory syncytial virus (RSV). For each day, we calculated the proportion of encounters related to ILI among all ED encounters. ILI counts were identified based on either positive laboratory test results or relevant ICD diagnosis codes. The participating EDs included: (1) Presbyterian University Hospital, (2) Children’s Hospital of Pittsburgh, (3) UPMC Shadyside, (4) McKeesport Hospital, and (5) UPMC Mercy Hospital. These five sites collectively represent a significant fraction (about 60%) of ED visits in Allegheny County and 30-40% of western PA. The research data were sourced from the University of Pittsburgh Neptune Research Data Warehouse (Visweswaran 2022).

The NRDM and ED datasets share strong geographic, and population overlap, as both focus on Allegheny County, Pennsylvania. We retrieved OTC sales data from 270 retail stores across the county, while the five EDs used for estimating prior disease probabilities also serve this region. Although the datasets are not individually linked, their shared regional focus supports the use of ED-based ILI daily incidence estimates in modeling NRDM purchasing patterns.

### 2.2 Model Development

The modeling used in this study focused on two classes of disease: (1) ILI and (2) OTHER, which is a broad category that includes all diseases other than ILI diseases. The main abbreviations used are summarized in Table 2.

**Table 2.** Main notation used in this paper.

| Symbols | Descriptions |
| --- | --- |
| $D$ | All OTC sales data |
| $D(p)$ | A single OTC product bought by a customer in purchase $p$; if a customer buys multiple products in one transaction, each product is treated as a separate purchase.<br>$D(p) \in \{\text{Cough or Cold, Antifever Adult, Antifever Pediatric, Baby or Child Electrolytes, Thermometers, Antidiarrheal, Hydrocortisones}\}$ |
| $pur_d$ | Total number of OTC purchases on day $d$ |
| $P_d(D)$ | Probability of the observed data on day $d$ |
| $P_d(D(p))$ | Probability of an individual purchase $p$ on day $d$ |
| $dx_i$ | Disease group $i$;<br>$dx_1 = \text{influenza – like illness (ILI)}; dx_2 = \text{diseases other than ILI (OTHER)}$ |
| $P_d(D(p), dx_i)$ | The joint probability that a customer with disease group $dx_i$ makes purchase $p$ on day $d$ |
| $P_d(D(p) \vert dx_i)$ | The conditional probability that a customer makes purchase $p$ on day $d$, given they have disease group $dx_i$ |
| $P_d(dx_i)$ | Prior probability that an OTC-purchasing customer has disease group $i$ |

For each day *d*, we used a probabilistic metric, *P*_*d*_(*D*), to identify the presence of unusual purchases on that day. This metric represents the probability of the observed data on day *d* (i.e., the number and types of OTC purchases on that day). We refer to this probability as the *daily likelihood*.

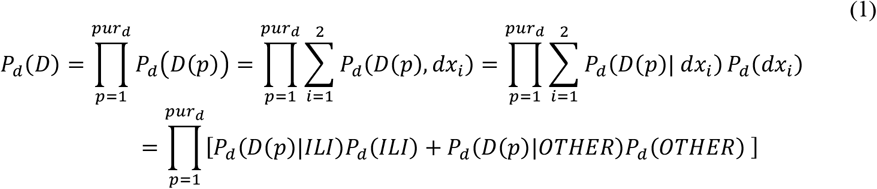

Equation 1 indicates how the daily likelihood *P*_*d*_(*D*) is calculated. We estimate this quantity by multiplying the probabilities of all individual purchase events *P*_*d*_(*D*(*p*)), assuming that these *pur*_*d*_ purchase events are independent. A purchase behavior typically reflects a customer’s (or their family member’s) illness; however, the specific underlying disease is not recorded in the NRDM database. Therefore, for each purchase behavior, we sum the joint probabilities, *P*_*d*_(*D*(*p*), *dx*_*i*_), across the disease categories, which in this paper are two disease groups.

Each joint probability is computed as the product of the prior probability of disease *dx*_*i*_ on day *d*, denoted as *P*_*d*_(*dx*_*i*_), and the conditional probability *P*_*d*_(*D*(*p*)| *dx*_*i*_), which is the probability of observing the purchase behavior given the presence of disease group *dx*_*i*_. To estimate these conditional probabilities in the model training period, we applied an expectation-maximization (EM) algorithm (see Section 2.3). <u>When training</u> the NRDM-Tracker model, we used the daily incidence of diseases in emergency departments in the same region on day *d*, as estimated by ILI Tracker, to estimate the prior probability, *P*_*d*_(*dx*_*i*_), as explained in Section 3.2. <u>When applying</u> the model to monitor for disease outbreaks, we used a uniform prior for *P*_*d*_(*dx*_*i*_), which does not require having access to ED data or to ILI Tracker.

### 2.3 Application of the Expectation-Maximization algorithm

Since many OTC products are used to relieve symptoms rather than treat specific diseases, we grouped conditions into two broad categories: ILI and OTHER. We applied the EM algorithm (see Algorithms 1 and 2 in the appendix) (McLachlan 2008) to estimate the conditional probabilities *P*_*d*_(*D*(*p*)| *dx*_*i*_), where *P*_*d*_(*D*(*p*) = *product*_*j*_| *dx*_*i*_ = *ILI* represents the probability a customer purchases product *j* on day *d* due to ILI either in that customer or in someone they are making the purchase (e.g., a family member). *P*_*d*_(*D*(*p*) = *product*_*j*_| *dx*_*i*_ = *OTHER* denotes the corresponding probability for patients with diseases other than ILI.

We used the ED data in the region in order to estimate the prior probabilities used by the EM algorithm that derives the conditional probabilities used in NRDM Tracker. To smooth the ED time series, we applied a centered 7-day moving average to the daily encounter counts. At the beginning and end of the series, where fewer neighboring values were available, a smaller window size (e.g., 4 or 5 days) was used to compute the average. This adaptive window ensures that smoothing can still be applied at the edges without discarding data. Based on the resulting smoothed counts, we calculated daily prior probabilities for ILI and OTHER that were used by the EM algorithm.

In applying EM to estimate the conditional probabilities above, we made the following assumptions:

1. **Time-invariance**: The conditional probabilities are constant across days, i.e., *P*_*d*_(*product*_*j*_ | *dx*_*i*_) is the same for all *d*. This assumes that given a disease-group category (e.g., ILI), the likelihood of purchasing a specific product remains stable over time. It is justified by the expectation that symptom-driven consumer behavior remains stable, especially when product availability and public health messaging are consistent. This assumption simplifies modeling by attributing temporal variation to disease-category daily incidence rather than to changes in individual purchasing behavior.
2. **Product narrowing**: We only monitored the purchase of the product categories shown in Table 1.

The EM algorithm (see Appendix for pseudocode) iteratively refines probability estimates through two main steps: the expectation step (E-step) and the maximization step (M-step). In the **E-step**, the algorithm computes the posterior probabilities *P*_*d*_(*ILI* | *product*_*j*_) and *P*_*d*_(*OTHER* |*product*_*j*_) for each product *j* on day *d*, using the current estimates of *P*G*product*_*j*_ | *ILI*) and *P*G*product*_*j*_ | *OTHER*), along with the prior probabilities of *ILI* and *OTHER* on day *d* estimated from ED data (see details in section 2.1).

In the **M-step**, the algorithm updates the conditional probabilities, *P*(*product*_)_ | *ILI*) and *P*(*product*_*j*_ | *OTHER*) by maximizing the expected likelihood. This is achieved by leveraging the weighted counts of product *j* purchases on day *d*, denoted as, 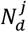, using the posterior probabilities obtained from the E-step. The E-step and M-step are repeated iteratively until convergence is achieved—that is, until the maximum change in probability estimates falls below a predefined threshold, *tol*.

### 2.4 Estimates of daily disease counts using NRDM data

After estimating the conditional probabilities *P*(*product*_*j*_ | *dx*_*i*_), we derive the expected number of customers with disease *dx*_*i*_ on day *d*, denoted *E*_*d*_(*dx*_*i*_), using Eq. 2. Specifically, *E*_*d*_(*dx*_*i*_) is calculated as the sum of the posterior probabilities *P*_*d*_(*dx*_*i*_ | *D*(*p*)) across all purchases on day *d*. For each individual purchase *p*, the posterior probability is computed via Bayes’ rule (Eq. 3), where the numerator is the joint probability, calculated as the product of the conditional probability of the purchase given the disease (as estimated via the EM algorithm) and the prior probability of the disease. The denominator normalizes this quantity by summing over the possible disease group categories, which in this paper is two categories.

In contrast to the EM training phase, where disease priors were learned from ED data, we fix the priors at *P*_*d*_(*ILI*) = 0.05 and *P*_*d*_(*OTHER*) = 0.95 for each day during the calculation with Eq. 3 to focus the contribution of NRDM data in estimating daily disease patterns. These values are consistent with the daily incidence observed in the ED data, where the daily *ILI* daily incidence ranged from 0 to 0.06 and the daily *OTHER* daily incidence ranged from 0.94 to 1 (Section 3.1, Figure 2).

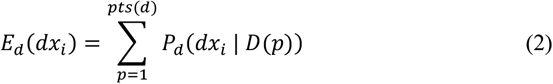

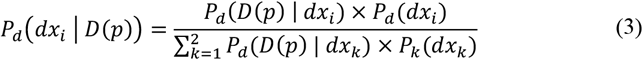

### 2.5 Detection of a distribution shift in relative product sales

Eq. 1 derives the probability (likelihood) of all the OTC purchases on day *d*; we will use *P*_*d*_(*D*) as shorthand to denote it. We also derive the following **adjusted likelihood**, which is the **geometric mean** of the daily likelihood across all purchases on day *d* (Eq. 4):

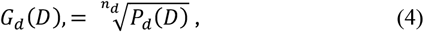

where *n*_*d*_ is the total number of the products purchased in the seven OTC categories on day *d*. The adjusted likelihood reflects the change in the distribution of the product-category purchases, whereas the original (unadjusted) likelihood is also sensitive to the number of product-category purchases per day.

### 2.6 Detection of a putative outbreak of a novel disease

We further analyzed the daily sales data and performed a statistical analysis to investigate abnormal patterns of sales that could indicate an outbreak of a novel disease. For each product *j*, we obtain the daily sales count 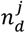 for day *d* and compute a right-sided p-value based on the empirical distribution of daily sales during a control period from day *d*-38 to day *d*-8, assuming a normal distribution. Approximately 68.8% (4326 out of 6284) of Shapiro–Wilk normality tests (Shapiro, 1965) passed the normality check, indicating that most control period row count distributions were consistent with a normal distribution.

Since a p-value is calculated for each day, over many monitored days, we applied the Benjamini–Hochberg procedure (Benjamini, 1995) to control the false discovery rate (FDR) at a significance level of 0.05. Specifically, we used the multiple-tests function from the Python statsmodels.stats.multitest module (statsmodels 0.15.0, 2024) with the method=‘fdr_bh’ option to correct for multiple hypothesis testing. This method adjusts raw p-values by scaling each value according to its rank and the total number of tests *m* (Eq. 5: where *m* is 6284 in our experiment; *p*_*j*_ are raw p-values; and the term *j* is the rank of the p-value in ascending order). A reverse cumulative minimum is then applied to ensure monotonicity across the adjusted p-values (Eq. 6), as described by Dudoit et al. (2003) and implemented in the Statsmodels package. These steps ensure that any day with an adjusted p-value at or below 0.05 is considered a statistically significant anomaly, while the expected proportion of false discoveries is controlled. Importantly, the adjusted p-values, *q*_*k*_, are guaranteed to be non-decreasing with respect to the ordered raw p-values.

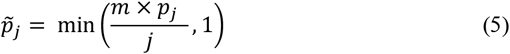

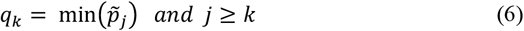

## 3. Results

This section presents the performance and behavior of the NRDM-based surveillance system. We begin by summarizing descriptive statistics of daily OTC product sales across seven symptom-related categories (Section 3.1). Next, we describe the estimation of model parameters using the EM algorithm (Section 3.2). We then evaluate the model’s ability to detect population-level anomalies during the COVID-19 pandemic using NRDM signals alone (Section 3.3). Finally, we assess the statistical significance of daily deviations in product sales across individual categories (Section 3.4).

### 3.1 Descriptive Statistics

Table 3 lists descriptive statistics of the daily counts of the seven categories from 274 stores in Allegheny County, PA from June 1, 2016 to December 30, 2021. The coverage of the NRDM for the Pittsburgh metropolitan statistical area, including Allegheny County, is 73% of all retail stores in the area. The median of the daily OTC sales count over the seven categories is 4062, with a wide range from 259 to 15,979, indicating substantial daily variation. The Cough or Cold category has the highest average daily sales (3,168), accounting for most of the total OTC purchases recorded in NRDM. Moderate-sale categories include Antifever for Adult, Baby or Child Electrolytes, and Antidiarrheal. Antifever for Pediatric, Hydrocortisones, and Thermometers have relatively low average sales. Most categories show right-skewed distributions (mean > median), suggesting occasional sales surges. The large variances (especially in Total and Cough or Cold categories) indicate irregular and potentially seasonal demand.

**Table 3.**
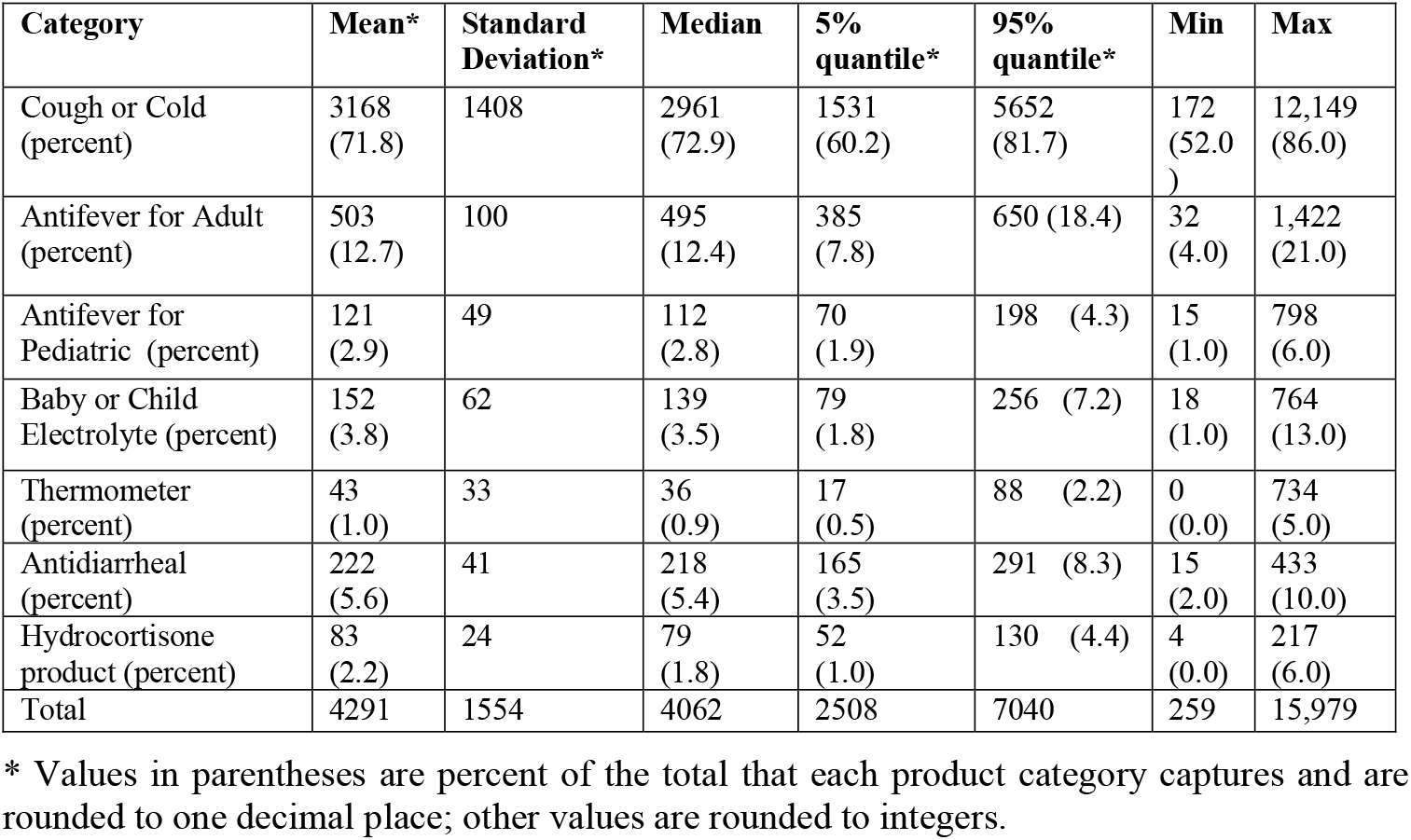
Summative statistics of daily sales of OTC categories from June 1, 2016 to December 30, 2021.

Figure 1 illustrates seasonal patterns in OTC product sales over five years. Both Total and Cough or Cold categories exhibit strong seasonal patterns, with recurring peaks during the winter months, reflecting the influenza and cold seasons. A prominent spike in early 2020 likely corresponds to the onset of COVID-19, followed by a dip and gradual return to seasonal trends.

**Figure 1.**
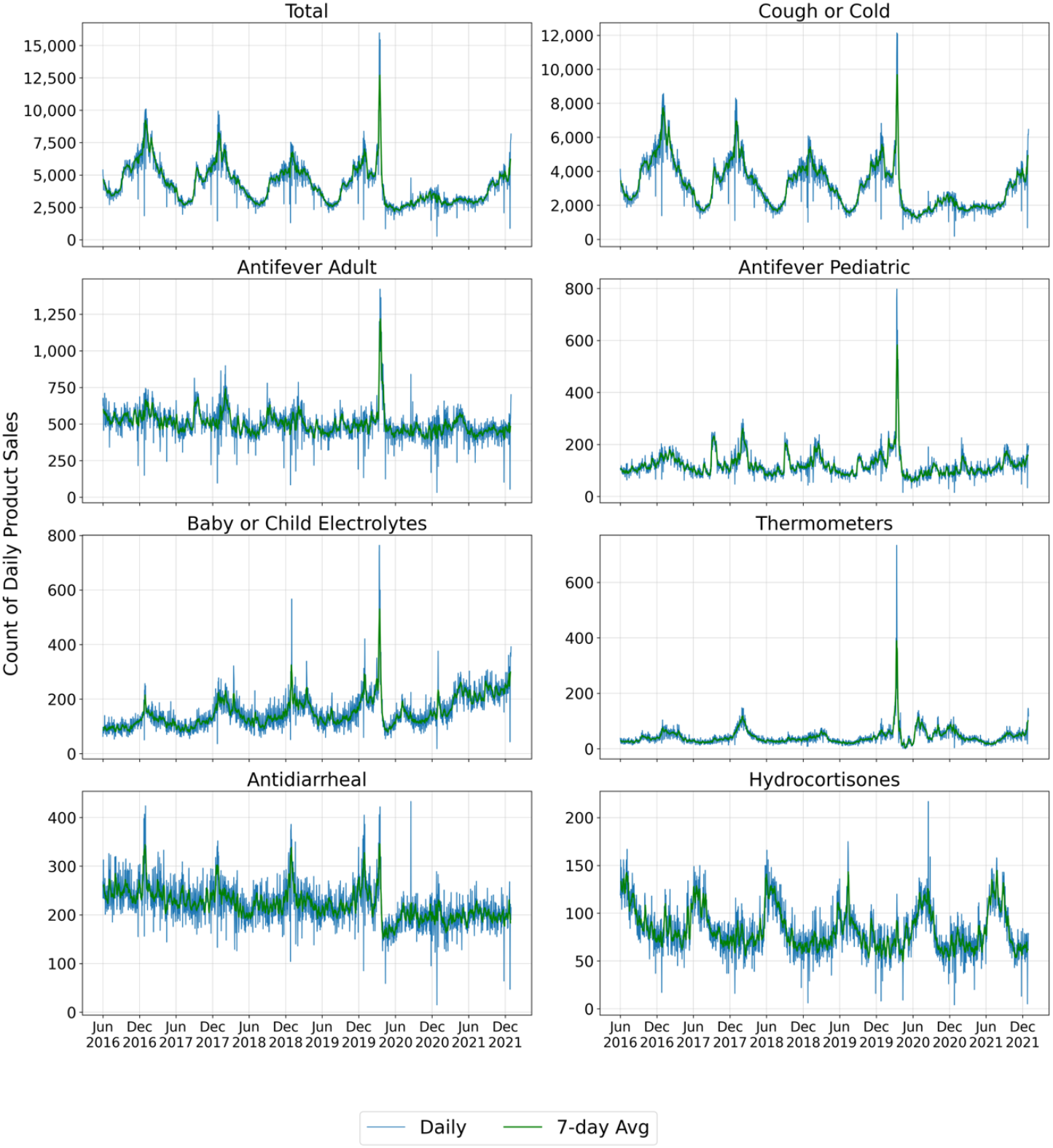
Temporal trends of daily product sales recorded in NRDM (the scales on the y axis differ across the plots).

**Figure 2.**
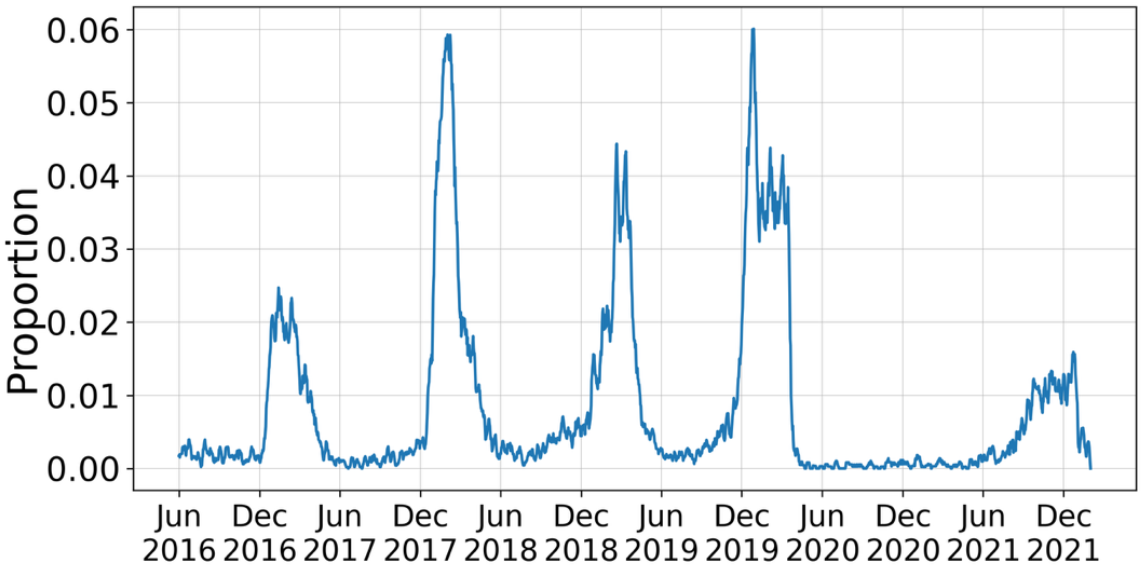
Temporal trends of daily disease priors estimated using ED data.

Sales trends among symptom-relief categories reveal varying degrees of seasonality and pandemic impact. Among antifever products, Adult Antifever shows relatively stable daily sales with modest fluctuations until a sharp surge in 2020, a pattern likely explained by the dual role of acetaminophen and ibuprofen as both antipyretic and analgesic medications, which contributes to their steady baseline demand. Pediatric Antifever follows a similar seasonal trend but at a lower scale. Cough or Cold products dominate in both volume and seasonality. Baby/Child Electrolytes show periodic increases with peaks in the winter. Antidiarrheal sales exhibit moderate daily variation with peaks around December. The peaks in Hydrocortisone sales are less pronounced but show a consistent seasonal pattern, with increased usage in warmer months, perhaps due to higher incidence of skin irritation, insect bites, and sun-related conditions. Diagnostic products display irregular or event-driven trends. For example, thermometer sales remain low at baseline but exhibit dramatic, event-driven spikes— particularly in early 2020—likely due to heightened public concern during the onset of the COVID-19 pandemic.

Figure 2 illustrates the temporal trends of these estimated priors. The left panel displays the estimated daily prior probability of ILI. The series shows clear seasonal peaks occurring annually—typically during the winter months (e.g., early 2017, early 2018, spring 2019, and early 2020)—consistent with typical seasonal trends of respiratory viruses in this region. A pronounced decline is observed in 2020 and 2021, reflecting reduced ILI-related emergency department visits during the initial phase of the COVID-19 pandemic in the U.S. A modest resurgence is seen in late 2021, coinciding with the return of seasonal respiratory virus activity.

### 3.2 Model Parameters

We divided the research data into training (June 1, 2016 – May 31, 2019) and test data (June 1, 2019 – December 30, 2021). Training data were used for model-parameter estimation. Test data were used for model performance evaluation and abnormal event detection.

Using Algorithm 2 (see the appendix), we estimated the conditional probabilities *P*(*product*_*j*_ | *ILI*) and *P*(*product*_*j*_ | *OTHER*). The EM algorithm converged after 36 iterations. Table 4 presents the estimated conditional probabilities, indicating the likelihood that each product category is purchased given an ILI–related or non-ILI-related encounter. These values reflect how specific each product is to ILI activity. Cough or Cold medications show a notably higher probability under ILI (0.792) than under OTHER (0.695), suggesting strong relevance to ILI-related symptoms. In contrast, Antidiarrheal and Hydrocortisones have lower probabilities under ILI (0.043 and 0.013, respectively) compared to OTHER (0.064 and 0.029), indicating they are more likely to be associated with non-ILI conditions. Antifever medications and electrolyte solutions show modest probabilities under both ILI and OTHER groups, which is consistent with their broader clinical use in treating fever and dehydration across a variety of types of illness along with antifever medications being used for pain treatment. These conditional probabilities are central to inferring ILI activity from purchase patterns and help distinguish symptom-specific products from those with more general therapeutic use.

**Table 4.** Conditional probabilities estimated by the EM algorithm.

| Category | P(product<sub>j</sub>\|ILI) | P(product<sub>j</sub>\|OTHER) |
| --- | --- | --- |
| Cough or Cold | 0.792 | 0.695 |
| Antifever for Adult | 0.095 | 0.142 |
| Antifever for Pediatric | 0.024 | 0.031 |
| Baby or Child Electrolyte | 0.026 | 0.032 |
| Thermometer | 0.008 | 0.008 |
| Antidiarrheal | 0.043 | 0.064 |
| Hydrocortisone | 0.013 | 0.029 |

\* Values are rounded to three significant digits.

### 3.3 Detection of Covid-19

Using Equations 2 and 3, NRDM Tracker calculated the expected counts of ILI from the NRDM data for the period from June 1, 2019 to December 30, 2021. Figure 3 illustrates the moving averages of the those expected ILI counts in blue. The yellow curve shows ILI counts derived from the ED cases for the following viruses: adenovirus, enterovirus, influenza, HMPV, PIV, and RSV. A disease caused by one of these viruses was considered present if a patient had a positive laboratory test result for the virus (e.g., a positive PCR test for influenza) or an ICD code indicative of the infection (listed in Table 5 in the Appendix). These <u>ED-based ILI tests and diagnoses excluded COVID-19 cases</u> which emulates the situation in which a novel disease may occur before it is explicitly identified and tested for. The correlation coefficient between two time series shown in blue and yellow is 0.66.

**Table 5.**
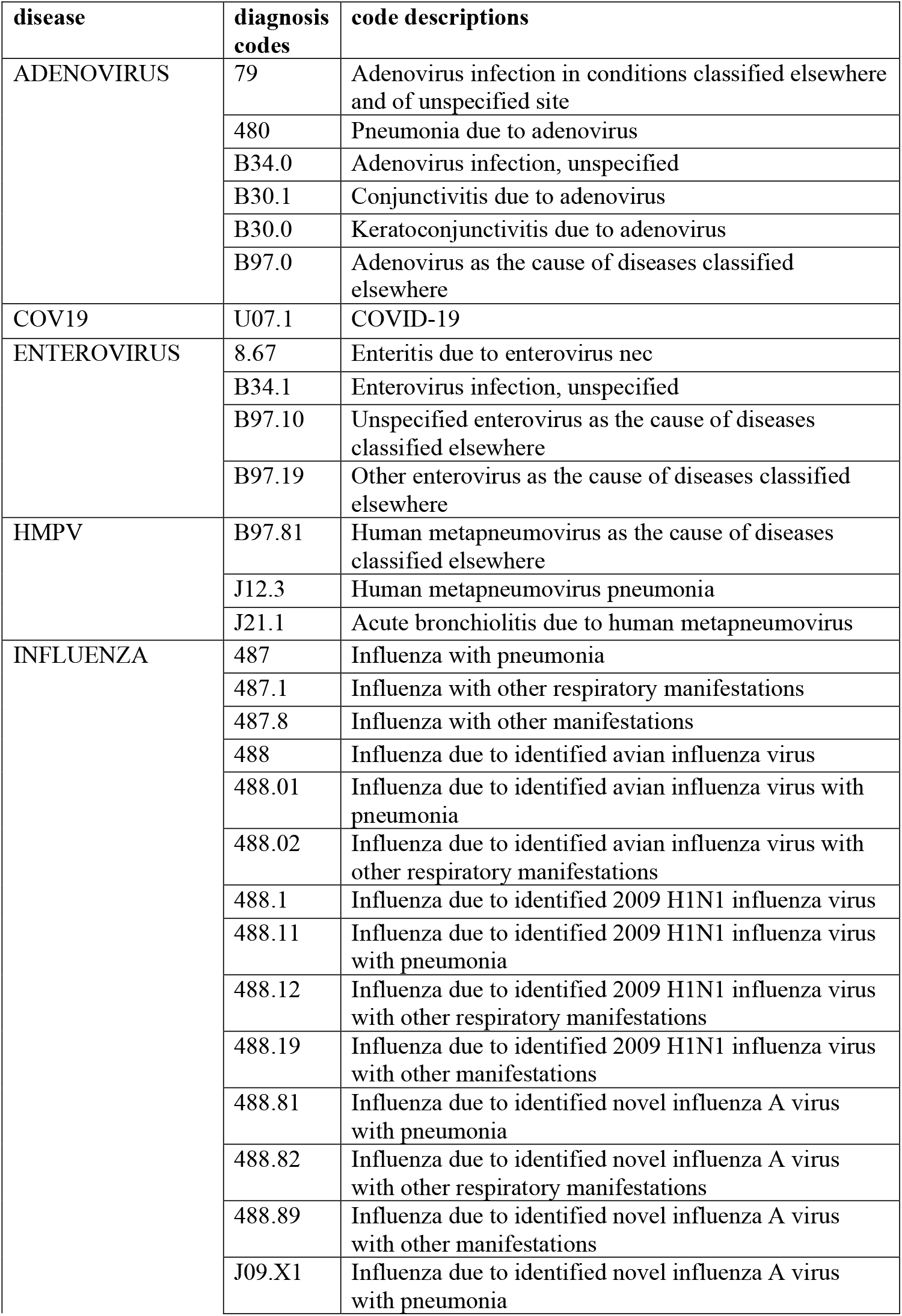
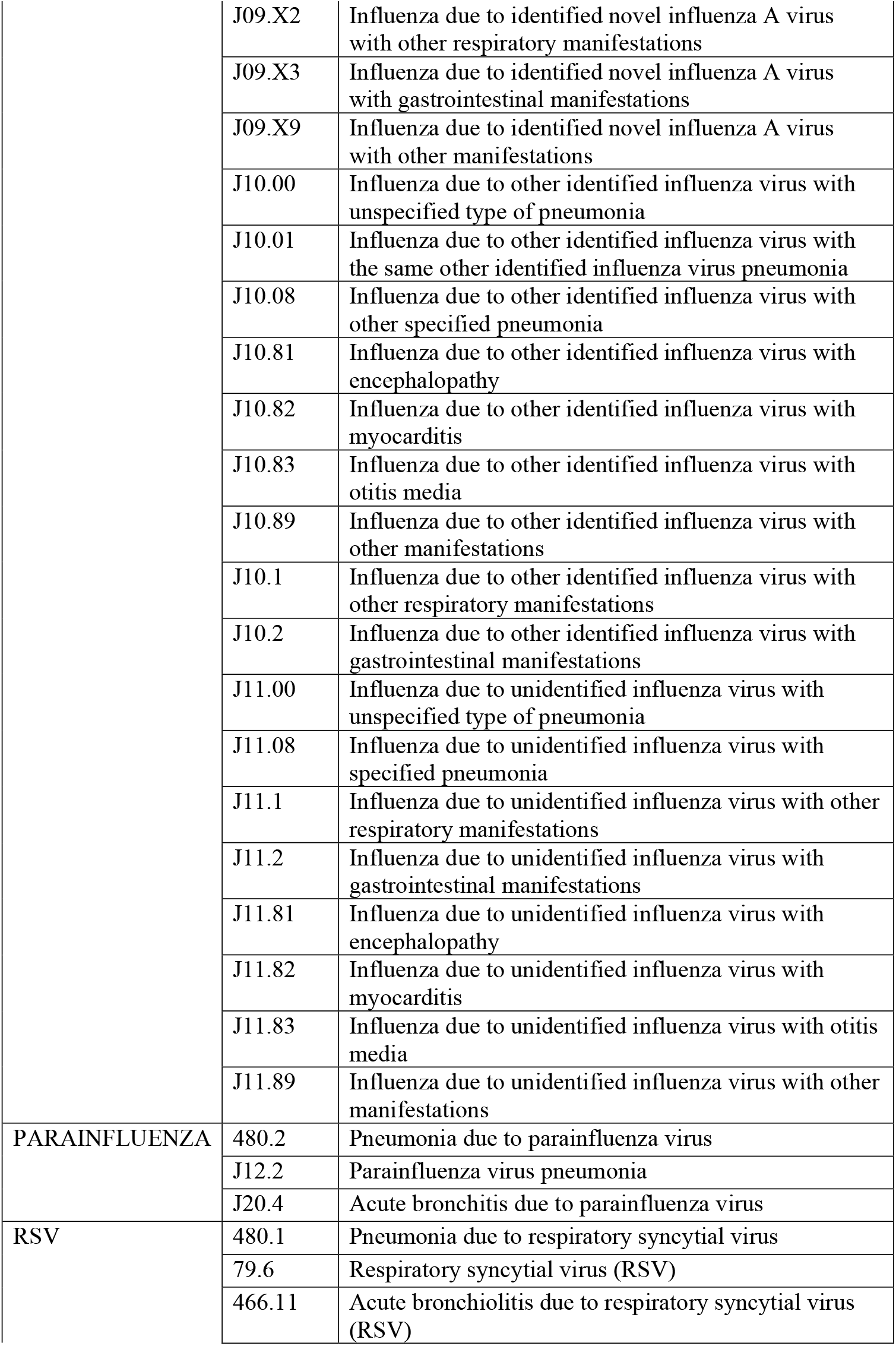
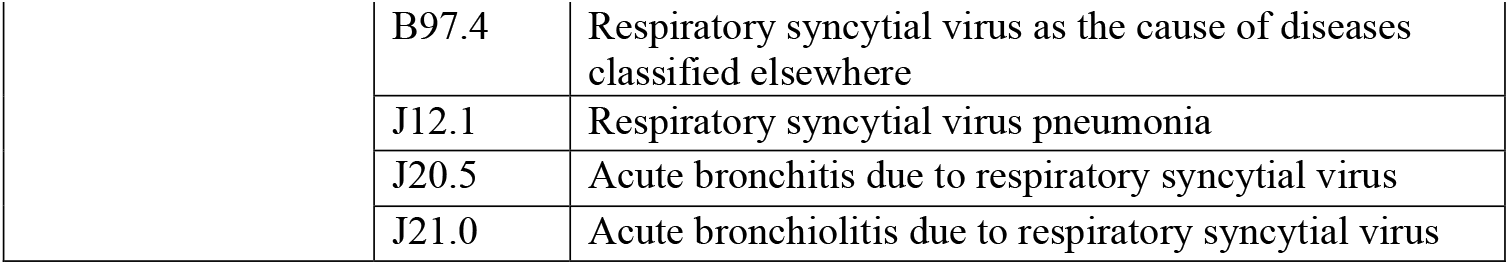
ICD codes related to influenza-like-illness.

| **disease** | **diagnosis codes** | **code descriptions** |
| --- | --- | --- |
| ADENOVIRUS | 79 | Adenovirus infection in conditions classified elsewhere and of unspecified site |
|  | 480 | Pneumonia due to adenovirus |
|  | B34.0 | Adenovirus infection, unspecified |
|  | B30.1 | Conjunctivitis due to adenovirus |
|  | B30.0 | Keratoconjunctivitis due to adenovirus |
|  | B97.0 | Adenovirus as the cause of diseases classified elsewhere |
| COV19 | U07.1 | COVID-19 |
| ENTEROVIRUS | 8.67 | Enteritis due to enterovirus nec |
|  | B34.1 | Enterovirus infection, unspecified |
|  | B97.10 | Unspecified enterovirus as the cause of diseases classified elsewhere |
|  | B97.19 | Other enterovirus as the cause of diseases classified elsewhere |
| HMPV | B97.81 | Human metapneumovirus as the cause of diseases classified elsewhere |
|  | J12.3 | Human metapneumovirus pneumonia |
|  | J21.1 | Acute bronchiolitis due to human metapneumovirus |
| INFLUENZA | 487 | Influenza with pneumonia |
|  | 487.1 | Influenza with other respiratory manifestations |
|  | 487.8 | Influenza with other manifestations |
|  | 488 | Influenza due to identified avian influenza virus |
|  | 488.01 | Influenza due to identified avian influenza virus with pneumonia |
|  | 488.02 | Influenza due to identified avian influenza virus with other respiratory manifestations |
|  | 488.1 | Influenza due to identified 2009 H1N1 influenza virus |
|  | 488.11 | Influenza due to identified 2009 H1N1 influenza virus with pneumonia |
|  | 488.12 | Influenza due to identified 2009 H1N1 influenza virus with other respiratory manifestations |
|  | 488.19 | Influenza due to identified 2009 H1N1 influenza virus with other manifestations |
|  | 488.81 | Influenza due to identified novel influenza A virus with pneumonia |
|  | 488.82 | Influenza due to identified novel influenza A virus with other respiratory manifestations |
|  | 488.89 | Influenza due to identified novel influenza A virus with other manifestations |
|  | J09.X1 | Influenza due to identified novel influenza A virus with pneumonia |
|  | J09.X2 | Influenza due to identified novel influenza A virus with other respiratory manifestations |
|  | J09.X3 | Influenza due to identified novel influenza A virus with gastrointestinal manifestations |
|  | J09.X9 | Influenza due to identified novel influenza A virus with other manifestations |
|  | J10.00 | Influenza due to other identified influenza virus with unspecified type of pneumonia |
|  | J10.01 | Influenza due to other identified influenza virus with the same other identified influenza virus pneumonia |
|  | J10.08 | Influenza due to other identified influenza virus with other specified pneumonia |
|  | J10.81 | Influenza due to other identified influenza virus with encephalopathy |
|  | J10.82 | Influenza due to other identified influenza virus with myocarditis |
|  | J10.83 | Influenza due to other identified influenza virus with otitis media |
|  | J10.89 | Influenza due to other identified influenza virus with other manifestations |
|  | J10.1 | Influenza due to other identified influenza virus with other respiratory manifestations |
|  | J10.2 | Influenza due to other identified influenza virus with gastrointestinal manifestations |
|  | J11.00 | Influenza due to unidentified influenza virus with unspecified type of pneumonia |
|  | J11.08 | Influenza due to unidentified influenza virus with specified pneumonia |
|  | J11.1 | Influenza due to unidentified influenza virus with other respiratory manifestations |
|  | J11.2 | Influenza due to unidentified influenza virus with gastrointestinal manifestations |
|  | J11.81 | Influenza due to unidentified influenza virus with encephalopathy |
|  | J11.82 | Influenza due to unidentified influenza virus with myocarditis |
|  | J11.83 | Influenza due to unidentified influenza virus with otitis media |
|  | J11.89 | Influenza due to unidentified influenza virus with other manifestations |
| PARAINFLUENZA | 480.2 | Pneumonia due to parainfluenza virus |
|  | J12.2 | Parainfluenza virus pneumonia |
|  | J20.4 | Acute bronchitis due to parainfluenza virus |
| RSV | 480.1 | Pneumonia due to respiratory syncytial virus |
|  | 79.6 | Respiratory syncytial virus (RSV) |
|  | 466.11 | Acute bronchiolitis due to respiratory syncytial virus (RSV) |

|  |  |
| --- | --- |
| B97.4 | Respiratory syncytial virus as the cause of diseases classified elsewhere |
| J12.1 | Respiratory syncytial virus pneumonia |
| J20.5 | Acute bronchitis due to respiratory syncytial virus |
| J21.0 | Acute bronchiolitis due to respiratory syncytial virus |

**Figure 3.**
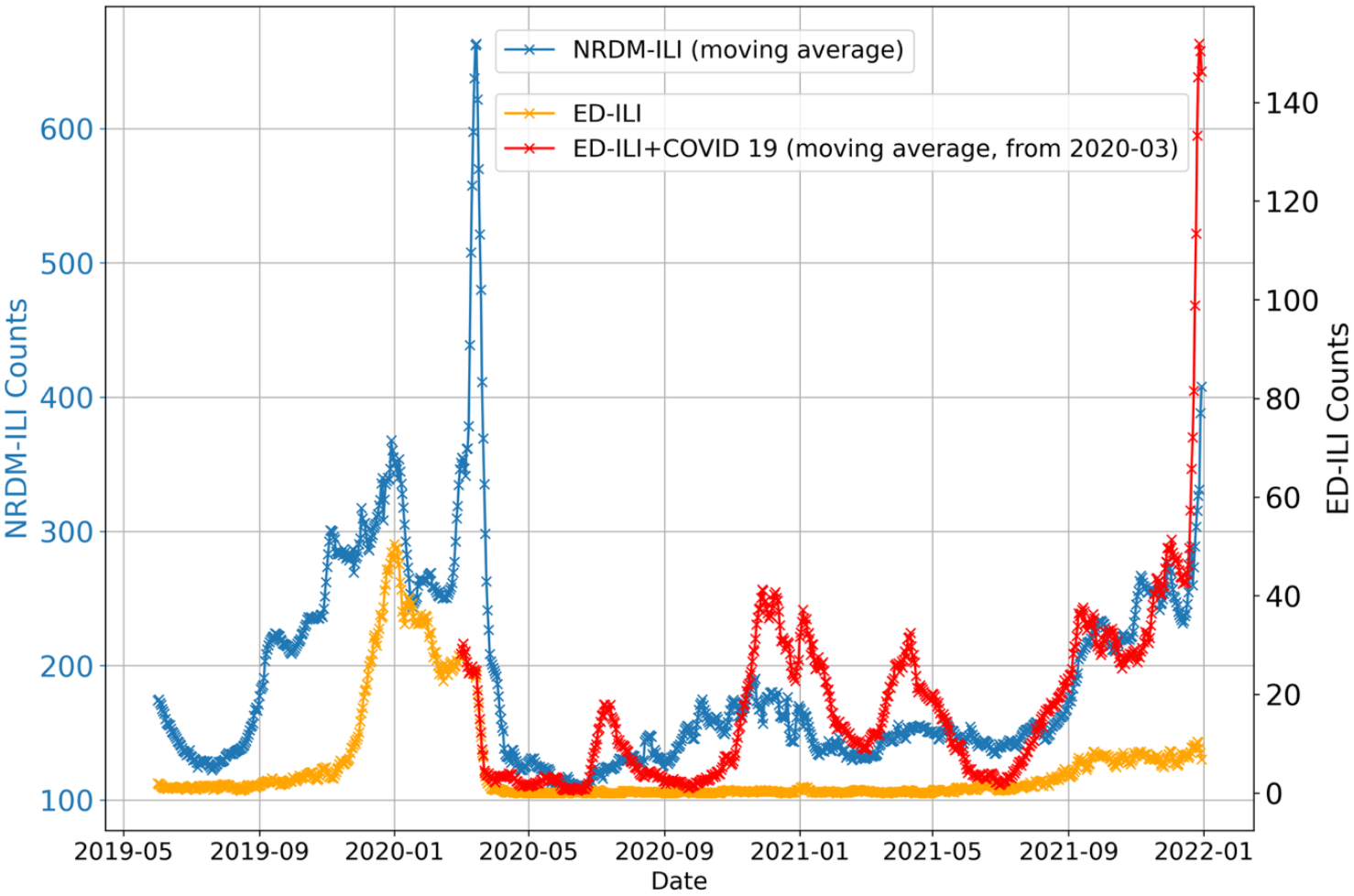
NRDM-tracker estimated ILI counts (blue) compared with ED-derived counts based on positive lab results and relevant ICD codes for a set of ILI diseases (yellow; see text), and with ED-derived counts including both ILI and COVID-19 cases (red).

The peak day of the NRDM-estimated ILI counts is March 13, 2020 (based on the underlying data, though not perfectly aligned with the moving average curve in Figure 3), coinciding closely with the clinical onset of the COVID-19 in Allegheny County. Because NRDM-ILI reflects ILI activity inferred from consumer purchasing behavior, the March 13th peak is likely due to precautionary purchasing triggered by widespread media coverage and growing public concern about COVID-19. The ILI counts from ED data have a peak on January 4, 2020 (not exactly aligned with the moving average curve). Both the blue and yellow curves in Figure 3 show an increase in late 2019 and very early 2020, corresponding to increased influenza and RSV activity observed in our ED data.

In addition, Figure 3 includes a red curve representing the ED-derived counts of ILI <u>and COVID-19</u> beginning in March 2020 which is when COVID-19 testing started in Allegheny County. As COVID-19 subsequently spread throughout the county, it became the dominant respiratory infectious agent, while influenza and RSV cases declined sharply. Consequently, the NRDM-ILI trend (blue curve) began to align more closely with the ED-derived counts of that includes COVID-19 (red curve), rather than the ED counts that do not (yellow curve).

Figure 4 contains two plots displaying the daily log likelihood of the OTC data computed by NRDM Tracker to monitor for unusual data patterns that may indicate the outbreak of a novel disease. The top panel shows the daily log likelihood. A prominent drop is visible around March 13, 2020. Interestingly, the top panel also shows sharp declines on other dates, such as July 2020, indicating statistically significant anomalies in purchasing volume. Notably, these events occurred even when the relative proportions of individual product categories did not change substantially (see below), implying that the signal was driven by spikes or drops in total purchase counts rather than a shift in product mix. The bottom panel depicts the adjusted daily log likelihood, computed as the geometric mean of category-level probabilities. This measure indicates whether there are changes in the relative proportions of product category purchases. The adjusted likelihood curve exhibits seasonal patterns and a sustained decline in mid-2020, followed by gradual recovery through 2021. The start of the COVID-19 outbreak does not appear to be associated with a strong change in the relative purchase of product categories.

**Figure 4.**
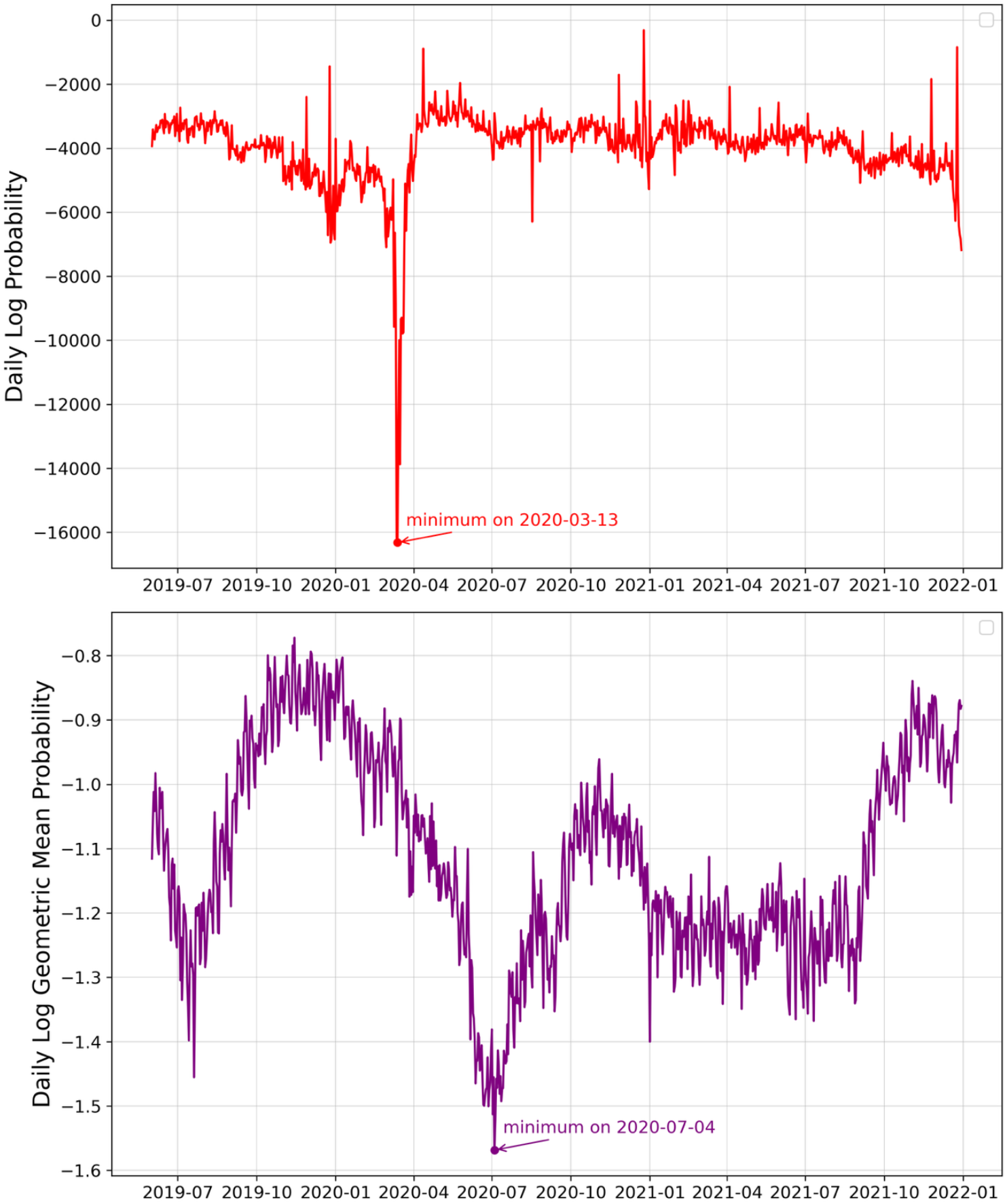
Daily log likelihood (top) and adjusted likelihood (bottom).

Using the Benjamini–Hochberg procedure, we computed the adjusted p-values for the daily counts of product sales across seven OTC categories: Antifever (Adult), Antifever (Pediatric), Baby or Child Electrolyte, Cough or Cold, Antidiarrheal, Hydrocortisone, and Thermometer. Figure 5 shows the time series of False-Discovery Rate adjusted p-values on a logarithmic scale using the Benjamini–Hochberg procedure (see Section 2.6), with the red dashed line indicating the significance threshold at p = 0.01. Sharp drops in p-values—particularly around March 2020—are visible across all categories, reflecting significant deviations in product purchasing behavior during the early phase of the COVID-19 pandemic. Several other spikes and dips are also observed in late 2020 and late 2021, corresponding to seasonal patterns or public health events.

**Figure 5.**
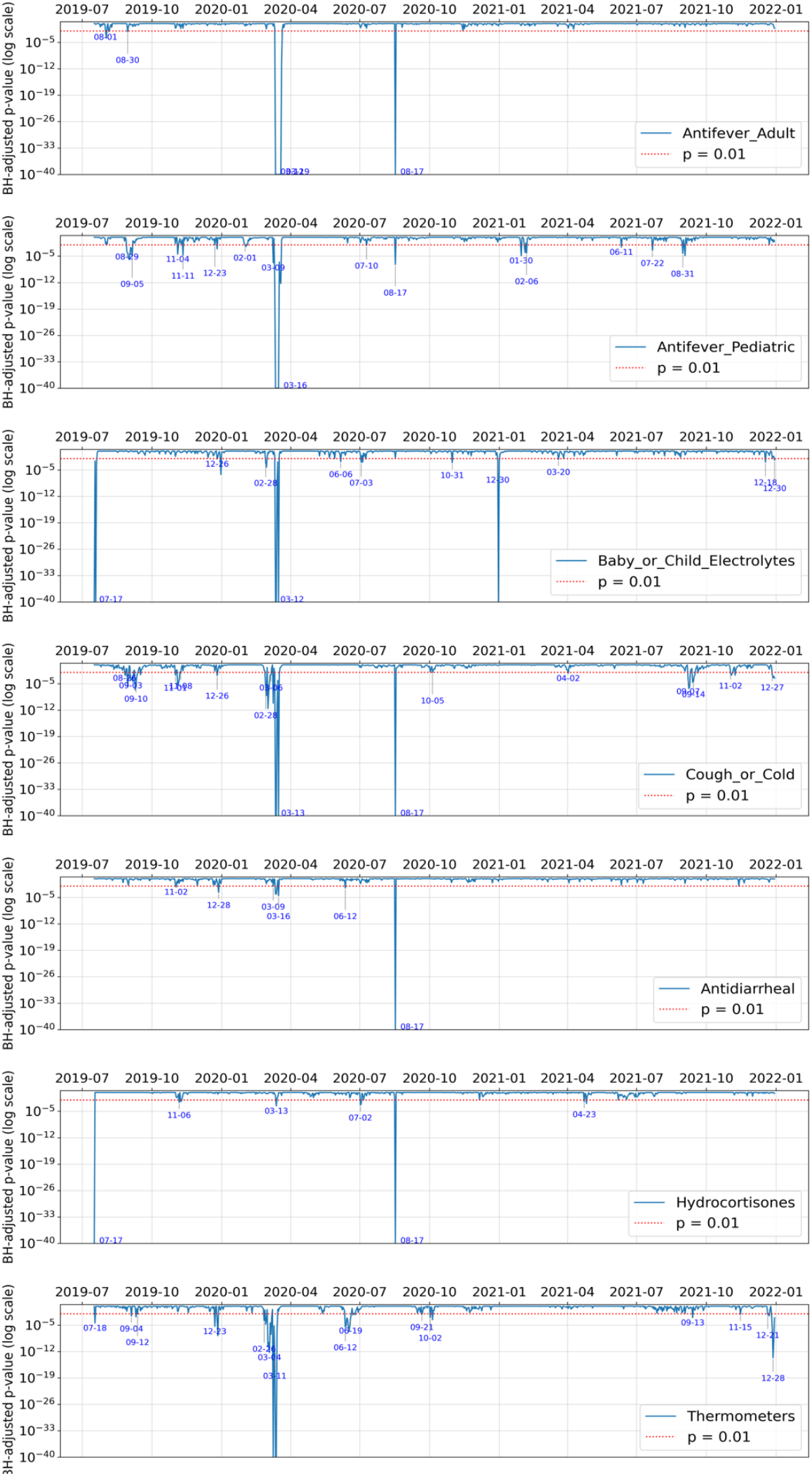
FDR-adjusted p-values for daily log likelihood.

## 4. Discussion

This paper demonstrates the potential for NRDM Tracker to be a useful tool for public health surveillance and early outbreak detection. By leveraging OTC product sales and applying a probabilistic modeling approach, we were able to estimate daily ILI activity and identify abnormal population-level purchasing patterns. The EM-based algorithm effectively estimated conditional probabilities that distinguish ILI-related purchasing behavior from that associated with other health conditions. The resulting ILI estimates from NRDM data exhibited moderately strong correlation (r = 0.66) with ILI-related ED visits, which provides support for the model’s alignment with measurable clinical rates of ILI disease in the region.

Our findings indicate that the NRDM-based ILI estimates, derived from OTC purchasing patterns, can capture public behavioral responses to disease outbreaks. These responses likely include purchases of cough and cold medications, antipyretics, and thermometers, even in the absence of corresponding clinical ILI encounters. Such dynamics could explain the divergence between the NRDM and ED curves in late 2019 and very early 2020, when NRDM estimates rose sharply while ED diagnoses for ILI remained comparatively stable. In general, while there is a close correspondence between the NRDM and ED curves in Figures 3, it is apparent that they are at times different, with an NRDM-derived estimate of increased ILI counts sometimes preceding the signal from ED data. Such patterns provide support that combining these two estimates of ILI activity could lead to better overall assessment of such activity in the population.

NRDM Tracker also derived daily likelihoods of observed sales patterns, and triggered alerts with frequentist hypothesis tests on daily log-likelihoods with Benjamini–Hochberg FDR control. The results demonstrated that likelihood-based signals can provide an early indication of abnormal health-related activity. The sharp drop in log-likelihood observed in March 2020 align well with the initial COVID-19 outbreak in the U.S.. Importantly, our right-tailed p-value framework—combined with multiple comparison correction—allowed for detection of statistically significant anomalies across multiple product categories. Our analysis also provides insights into the product sales that drive the detection of an anomaly.

There are limitations of our approach. First, NRDM sales data do not directly record patient diagnoses, making it challenging to validate findings against gold-standard clinical data without assumptions. Second, behavioral factors such as panic buying, supply chain disruptions, or public advisories may also influence OTC sales, potentially leading to false-positive signals about a new disease outbreak. Lastly, our modeling assumes stable conditional probabilities over time, which may not hold true under significant shifts in disease presentation or consumer behavior. This limitation could be attenuated by periodically updating the conditional probabilities. Taken together, these limitations point to remaining challenges in differentiating between behavior-driven anomalies and true disease-driven signals, adjusting for product substitution, and developing models that can adapt to evolving population health behaviors.

Future research may enhance this work by incorporating additional data sources (e.g., social media signals) or by applying adaptive models that account for evolving purchasing behaviors. Moreover, real-time implementation and evaluation in public health settings could evaluate the utility of the approach for early detection and decision support.

Despite its successful use for over two decades, the operators of the NRDM have decided to cease operations on December 31, 2025, due to the lack of stable funding support. Public health departments have found it increasingly difficult to obtain funds to subscribe to the service and retailers had become increasingly reluctant to share data. Nevertheless, the study reported here provides an assessment of the usefulness of such data, which may become available again in the future.

## 5. Conclusions

Our results suggest that NRDM-based data, when paired with probabilistic modeling and anomaly detection techniques, may provide an effective supplement to traditional disease monitoring systems. The observed alignment between NRDM-derived ILI estimates and ED-reported ILI cases, along with the detection of known public health events (e.g., COVID-19 onset), support the system’s potential for early detection of both seasonal and novel disease activity. By identifying significant deviations in symptom-related OTC product sales, the system could offer timely and actionable public-health insights. Continued refinement and integration with other data sources and modeling frameworks may further strengthen its role in the national biosurveillance ecosystem.

## Data Availability

All data produced in the present study are available upon reasonable request to the authors

## Acknowledgements

This research was supported by grant R01LM013509 (Automated Surveillance of Overlapping Outbreaks and New Outbreak Diseases) from the National Library of Medicine (NLM) of the U.S. National Institutes of Health (NIH). The research data from Emergency Departments were retrieved by the University of Pittsburgh Health Record Research Request, which was supported by the NIH grant UL1TR001857. Harry Hochheiser and Jessi Espino also received support from NIGMS grant U24GM132013 (MIDAS Coordination Center) and NIGMS grant R24GM153920 (MIDAS Coordination Center). Jessi Espino also received support from CDC grant 5U01IP001184 (Evaluating Respiratory Virus Vaccine Effectiveness in a Large, Diverse Healthcare System). Ye Ye also received support from NLM grant R00LM013383 (Transfer Learning to Improve the Re-usability of Computable Biomedical Knowledge). Marian G. Michaels also received support from CDC grant U01IP001152 (New Vaccine Surveillance Network).

## IRB Approval

The research protocol was approved by University of Pittsburgh IRB Study 20030193.

## Appendix

### Algorithm 1

Expectation-Maximization (EM) Algorithm

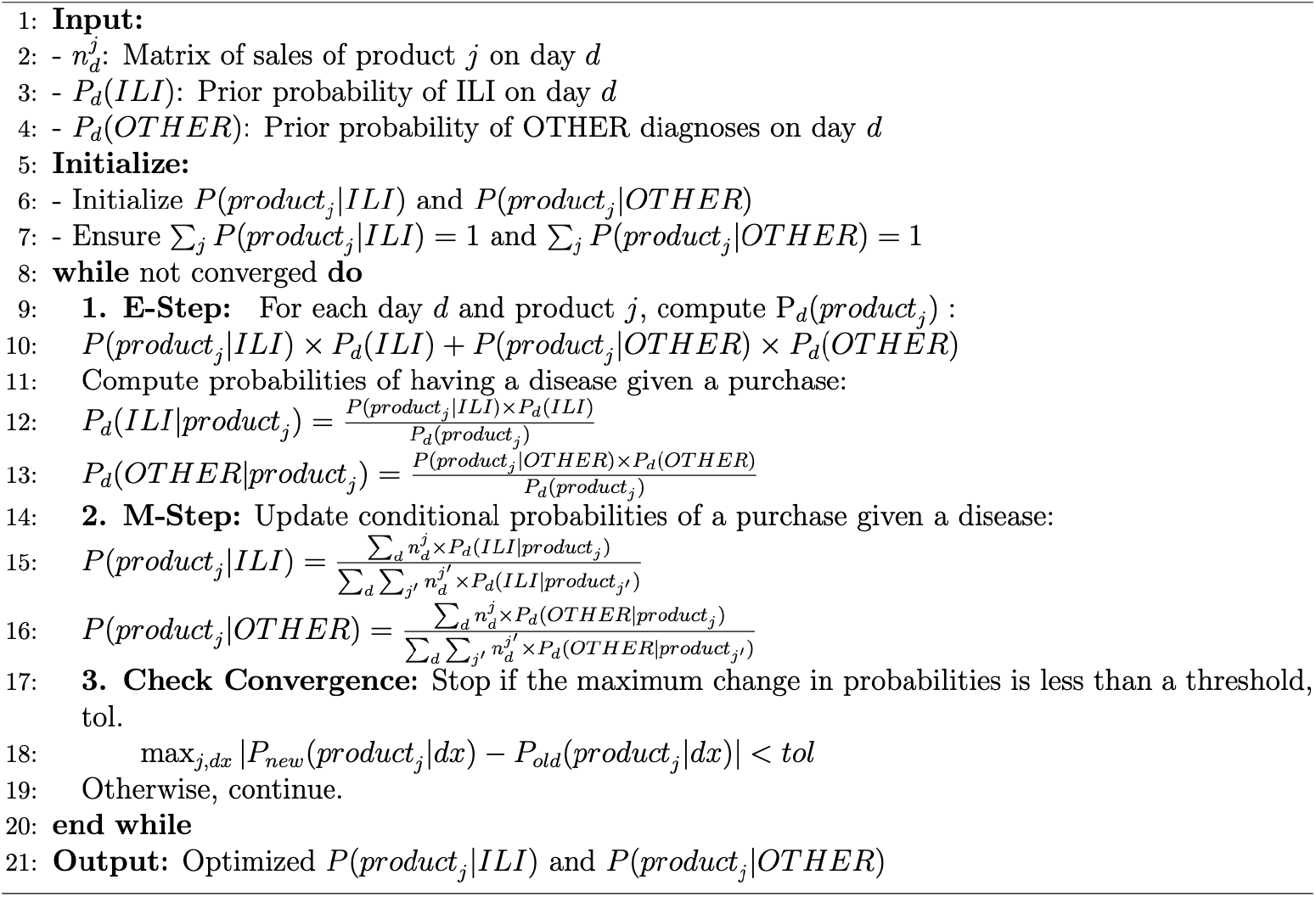

The robust implementation of the EM algorithm (Algorithm 2) extends Algorithm 1 by incorporating enhancements to improve numerical stability and convergence reliability. It refines the E-step by computing log-likelihoods and applying a stabilized posterior probability calculation, which mitigates issues related to numerical underflow. In the M-step, a smoothing constant ℰ is introduced to ensure stable updates of the conditional probabilities, thereby avoiding zero estimates in scenarios with sparse data. These modifications enhance the algorithm’s robustness and practical applicability while preserving the core iterative structure of Algorithm 1—alternating between updating posterior probabilities and refining conditional probability estimates until convergence is reached.

In our experiments, we initialize *P*(*product*_*j*_|*ILI*) using a uniform distribution across the seven products (i.e., each product is assigned a probability of 1/7). Similarly, *P*(*product*_*j*_|*OTHER*) is initialized uniformly across all products. We set the smoothing constant ϵ to 1×10^−8^, and the convergence threshold for probability changes, *tol*, is also set to 1×10^−8^.

### Algorithm 2

EM algorithm (Robust Implementation)

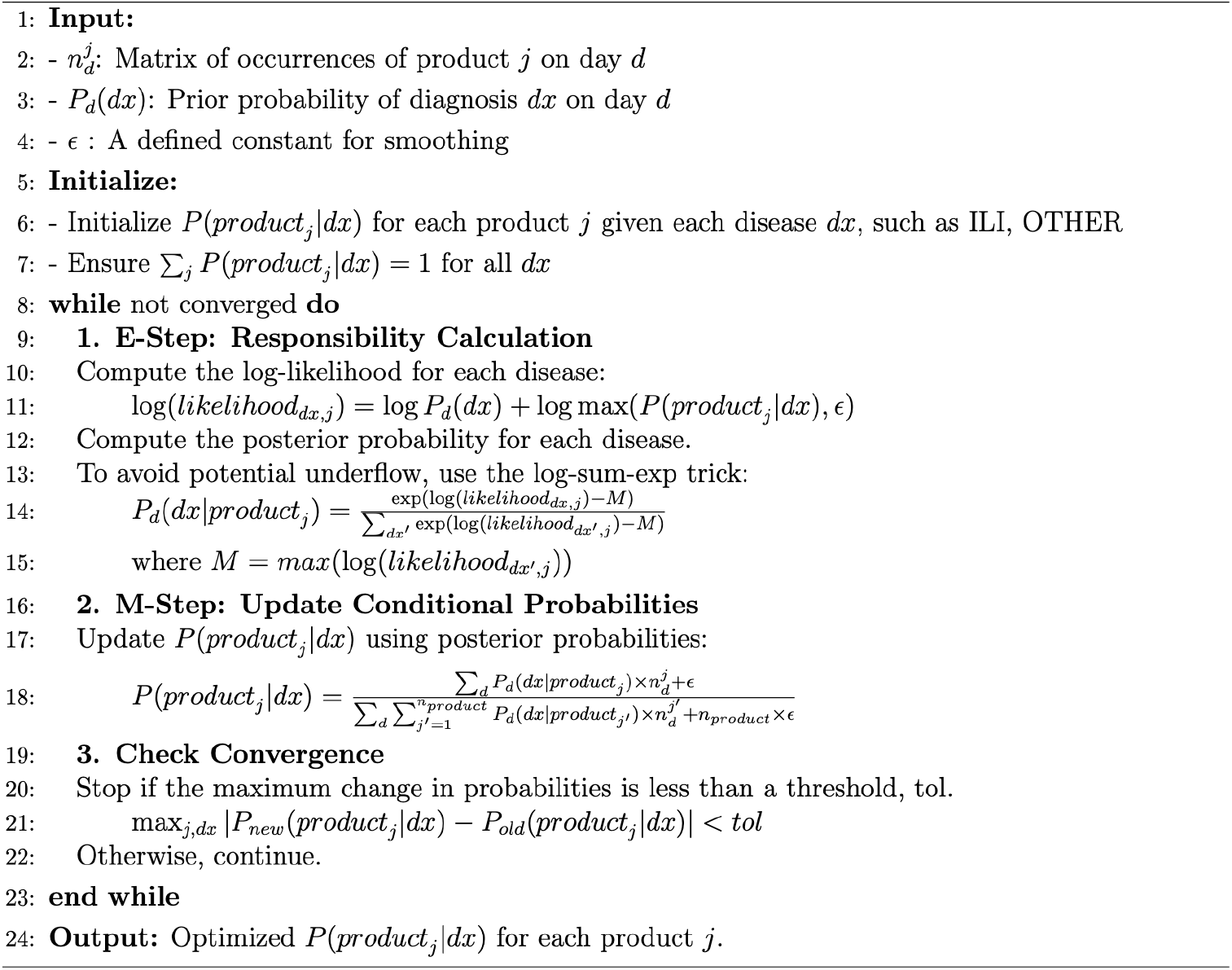

